# The impact of relaxing interventions on human contact patterns and SARS-CoV-2 transmission in China

**DOI:** 10.1101/2020.08.03.20167056

**Authors:** Juanjuan Zhang, Maria Litvinova, Yuxia Liang, Wen Zheng, Huilin Shi, Alessandro Vespignani, Cecile Viboud, Marco Ajelli, Hongjie Yu

**Affiliations:** School of Public Health, Fudan University, Key Laboratory of Public Health Safety, Ministry of Education, Shanghai, China; ISI Foundation, Turin, Italy; Laboratory for the Modeling of Biological and Socio-technical Systems, Northeastern University, Boston, MA USA; Division of International Epidemiology and Population Studies, Fogarty International Center, National Institutes of Health, Bethesda, MD, USA; Department of Epidemiology and Biostatistics, Indiana University School of Public Health, Bloomington, IN, USA

## Abstract

Non-pharmaceutical interventions to control COVID-19 spread have been implemented in several countries with different intensity, timing, and impact on transmission. As a result, post-lockdown COVID-19 dynamics are heterogenous and difficult to interpret. Here we describe a set of contact surveys performed in four Chinese cities (Wuhan, Shanghai, Shenzhen, and Changsha) during the pre-pandemic, lockdown, and post-lockdown period to quantify the transmission impact of relaxing interventions via changes in age-specific contact patterns. We estimate that the mean number of contacts increased 5%-17% since the end of the lockdown but are still 3-7 times lower than their pre-pandemic levels. We find that post-lockdown contact patterns in China are still sufficiently low to keep SARS-CoV-2 transmission under control. We also find that the impact of school interventions depends non-linearly on the share of other activities being resumed. When most community activities are halted, school closure leads to a 77% decrease in the reproductive number; in contrast, when social mixing outside of schools is at pre-pandemic level, school closure leads to a 5% reduction in transmission. Moving forward, to control COVID-19 spread without resorting to a lockdown, it will be key to dose relaxation in social mixing in the community and strengthen targeted interventions.

**One Sentence Summary:** Social contacts estimated in the post-lockdown period in four large Chinese cities are not sufficient to sustain local SARS-CoV-2 transmission.

## Introduction

The novel coronavirus disease 2019 (COVID-19) outbreak caused by SARS-CoV-2 began in Wuhan City, China in December 2019 and quickly became a global pandemic on March 11, 2020 (*1*). As of July 30, a total of 84,292 cases of COVID-19, including 4,634 deaths, have been reported in mainland China (*2*). The epidemic in Wuhan and in the rest of China subsided quickly after implementation of strict containment measures and travel restrictions. With the reduction of domestic cases in China and recent cases primarily originating from international travel(*2*), strict interventions have been gradually relaxed since February 2020.

In most of mainland China outside of Hubei Province, workplaces were allowed to resume operations after February 10, 2020 (*3*). Close community management (e.g., only one household member was allowed to purchase supplies every three days) and travel restrictions were lifted in March, and schools reopened at the end of April, 2020. Similarly, the lockdown ended on April 8, 2020 in Wuhan. It has been widely argued that relaxing inventions increases the risk of resurgence as transmission may intensify, a prediction that has been borne out in the Southern US and Europe (*4-8*). However, despite interventions being relaxed and continuous importation of new infections, a second wave in China has yet to materialize. Key questions remain about how relaxing interventions alters age-specific contact patterns and may in turn affect transmission. Quantifying the interaction between interventions and contacts is a key step to understand the post-lockdown SARS-CoV-2 transmission patterns.

This study aims to measure changes in mixing patterns as interventions are gradually relaxed to understand their impact on epidemic spread. To achieve this objective, we collected contact survey data in three different phases of the pandemic (before the pandemic, during the lockdown, and while interventions were being relaxed) in four locations in China (Wuhan, Shanghai, Shenzhen, Changsha). Based on the collected data, we investigate changes in age-stratified contact patterns and provide a model-based evaluation of their impact on the SARS-CoV-2 transmission. Moreover, we leverage the calibrated model to project the impact of a hypothetical further increase in the number of contacts on the emergence of a second wave.

## Results

### Contact surveys

We performed diary-based contact surveys(*9-12*) in four Chinese cities (Wuhan, Shanghai, Shenzhen and Changsha) representing different epidemiological situations. Wuhan was the early epicenter of the outbreak. Shanghai is a highly connected international hub and, thus, has been experiencing continuous importations of COVID-19 cases. Shenzhen is a major hub in Guangdong Province, which reported the largest number of COVID-19 cases outside Hubei Province. Changsha is a city of Hunan Province adjacent to Hubei Province with large number of commuters’ influx to and from Hubei.

The surveys were conducted from March 1 to March 20, 2020 in Shanghai, Shenzhen, and Changsha, about one month after workplaces started to reopen on February 10. In Wuhan the survey was performed from May 7 to May 15, 2020, about one month after the end of the lockdown (April 8). We collected information on weekday contact behavior for the pre-COVID-19 baseline period, the pandemic period when strict social distancing was in place, and during the post-lockdown survey period, as interventions were relaxed (Material and Methods). At the time of the contact survey, schools had not reopened except for the final year of senior high school in Wuhan, but travel restrictions and community management had begun to relax (Fig. 1) (Supplementary Material, Sec. 1).

**Fig. 1.**
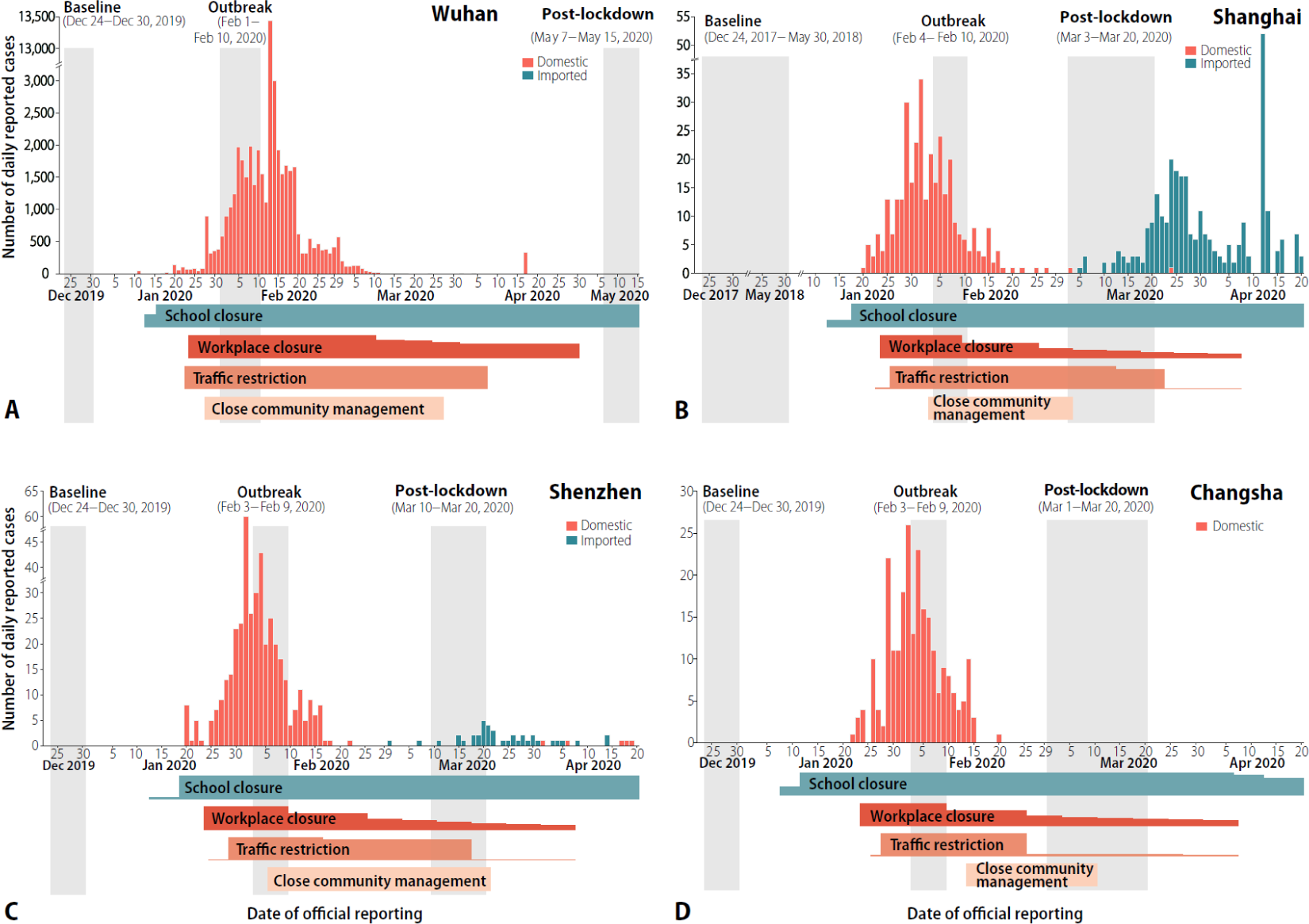
Number of COVID-19 reported cases, timing of the surveys, and the main interventions in place over time. (**A)** Wuhan. (**B)** Shanghai. (**C)** Shenzhen. (**D)** Changsha. COVID-19 cases by local transmission and international importation are indicated in light red and blue. The contact surveys for Shanghai, Shenzhen, and Changsha were conducted from March 1 to March 20, corresponding to the period of relaxation of interventions, and May 7 to May 15 for Wuhan. The contact diaries for the baseline and outbreak periods were derived from retrospective interviews or previous surveys. The change of four main interventions over time were put at the bottom of each panel, where the height of horizontal strips refers to the relative proportion of population affected by the intervention (up to 100%). Details on changed interventions are in Tab. S1 of the Supplemental Material.

### Characteristics of contact patterns

We analyzed a total of 54,324 contacts reported by 9,206 study participants (Table 1). In Wuhan, the average daily number of contacts per participant significantly increased from 2.0 during the outbreak period (mean contacts weighted by age structure: 1.9) to 3.3 in the post-lockdown period (mean contacts weighted by age structure: 3.6) (p<0.001). The increase in contacts was significantly different by sex, age, type of profession, and household size, with the exception of respondents under 20 years (Table 1). A smaller increase was observed in Shanghai, Shenzhen, and Changsha, where the average daily number of contacts increased by 0.1-0.4 contacts per day (0.2-0.8 for the mean number of contacts weighted by the age structure) starting from about 2.2 contacts during the lockdown period. The observed increase in the number of contacts (especially among adults) is statistically significant for Wuhan, Shanghai, and Shenzhen (Tab. 1). Participants in the four cities essentially reported the same number of contacts during the lockdown, but on average individuals in Wuhan reported more contacts than individuals in the other cities in the post-lockdown period, possibly due to the different timing of the contact surveys in the four locations (Supplementary Material, Sec.3.2).

**Table 1.**
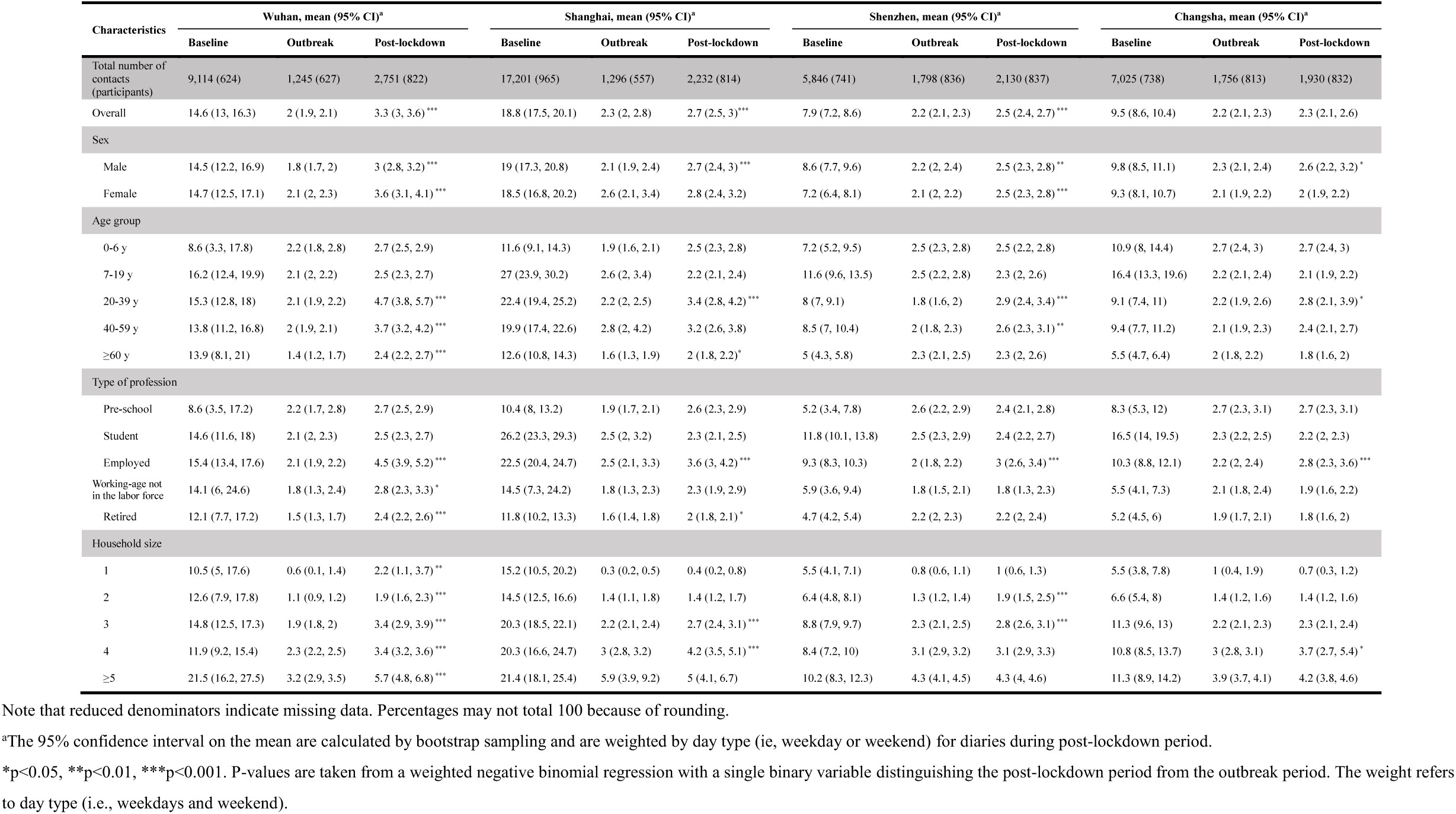
Number of contacts by respondent characteristics, location and time period.

In the four cities, the vast majority of contacts occurred at home in both periods, but workplace contacts increased in the post-lockdown period (Supplementary Material, Sec.3.3 and 3.4). Moreover, a statistically significant increase in the number of physical (skin-to-skin) contacts was also observed in Wuhan only, but the increase was relatively small – from 1.2 to 1.4 physical contacts (Supplementary Material, Sec.3.3). Overall, the number of contacts increased over time from the outbreak period to the post-lockdown period (p<0.001) (Supplementary Material, Sec.3.6).

When we consider the pre-pandemic (baseline) period, the typical features of age-mixing patterns(*10, 11*) emerge in all locations (Fig. 2A-2D). These features can be illustrated in the form of age-stratified contact matrices (provided as ready-to-use tables in the Supplementary Materials, Sec. 3.7), where each cell represents the mean number of contacts that an individual of a certain age group has with other individuals of different ages. The bottom left corner of the matrix, corresponding to contacts between school-age children, is where the largest number of contacts is recorded. The contribution of contacts in the workplace is visible in the central part of the matrix, while the three diagonals (from bottom left to top right) represent contacts between household members.

**Fig. 2.**
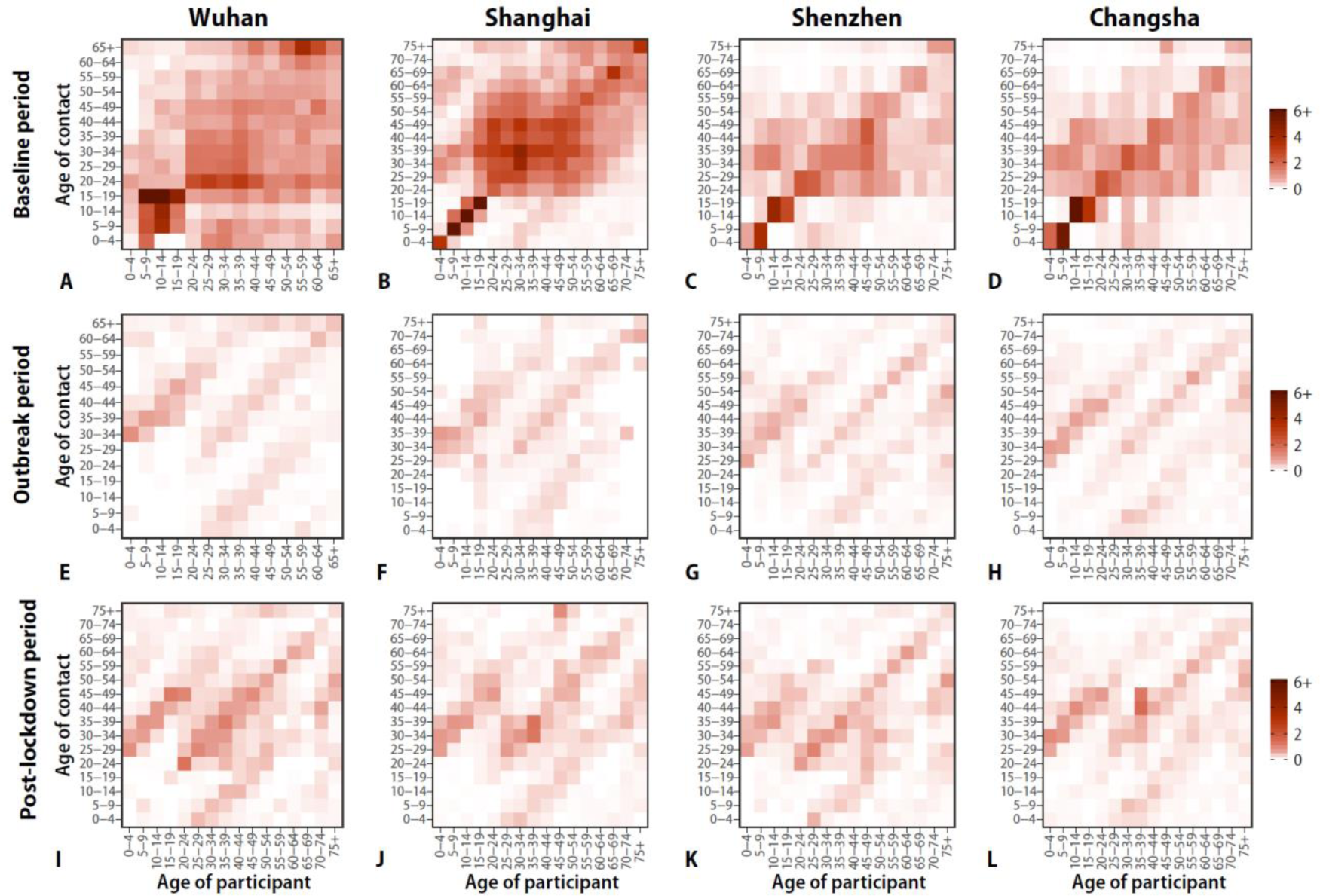
Contact matrices by age. (**A**) Baseline period contact matrix for Wuhan (regular weekday only). Each cell of the matrix represents the mean number of contacts that an individual in a given age group has with other individuals, stratified by age groups. The color intensity represents the numbers of contacts. To construct the matrix, we performed bootstrap sampling with replacement of survey participants weighted by the age distribution of the actual population of Wuhan. Every cell of the matrix represents an average over 100 bootstrapped realizations. (**B**) Same as (A), but the contact matrix weighted by weekday and weekends for Shanghai. (**C and D**) Same as (A), but for Shenzhen and Changsha. (**E-H**) Same as (A), but for the outbreak contact matrices for Wuhan, Shanghai, Shenzhen, and Changsha. (**I-L**) Same as (A), but for the post-lockdown contact matrices weighted by weekday and weekends for Wuhan, Shanghai, Shenzhen, and Changsha.

During the COVID-19 outbreak period, when strict social distancing policies were in place, most of the above-mentioned features disappear, essentially leaving the sole contribution of household mixing (Fig. 2E-2H). In particular, assortative contacts between school-age individuals fully vanish. Overall, contacts during the outbreak mostly occurred within household members (more than 90% in Shenzhen, Changsha and Wuhan; 79% in Shanghai). If we compare the post-lockdown period with the outbreak period, more contacts were reinstated in the workplace and in the community settings. Therefore, the fraction of contacts at home decreased to 63.0% in Wuhan, 72.5% in Shanghai, 70.2% in Shenzhen, and 78.8% in Changsha. Nevertheless, the three diagonals (from bottom left to top right) representing contacts between household members were still dominant. It is important to note that while strict social distancing measures gradually relaxed in these four locations, a considerable share of workers had not yet resumed work or continued to work from home at the time of the survey (Supplementary Material, Sec. 3.5).

### Modeling the impact of relaxing interventions on SARS-CoV-2 transmission

We used the next generation matrix approach to quantify changes in R_0_ during the post-lockdown period, as interventions were relaxed. In the early phases of COVID-19 spread in Wuhan, before control measures were put in place, R_0_ was estimated to range from 2.0 to 3.5(*13-19*). In this study, we extended this range from 1 to 4 for the baseline period (i.e., before interventions). We found that the considerable changes of mixing patterns observed in Shenzhen and Changsha during the social distancing period led to a drastic decrease in R_0_ (Fig. 3). The reproductive number drops well below the epidemic threshold for all four locations.

**Fig. 3.**
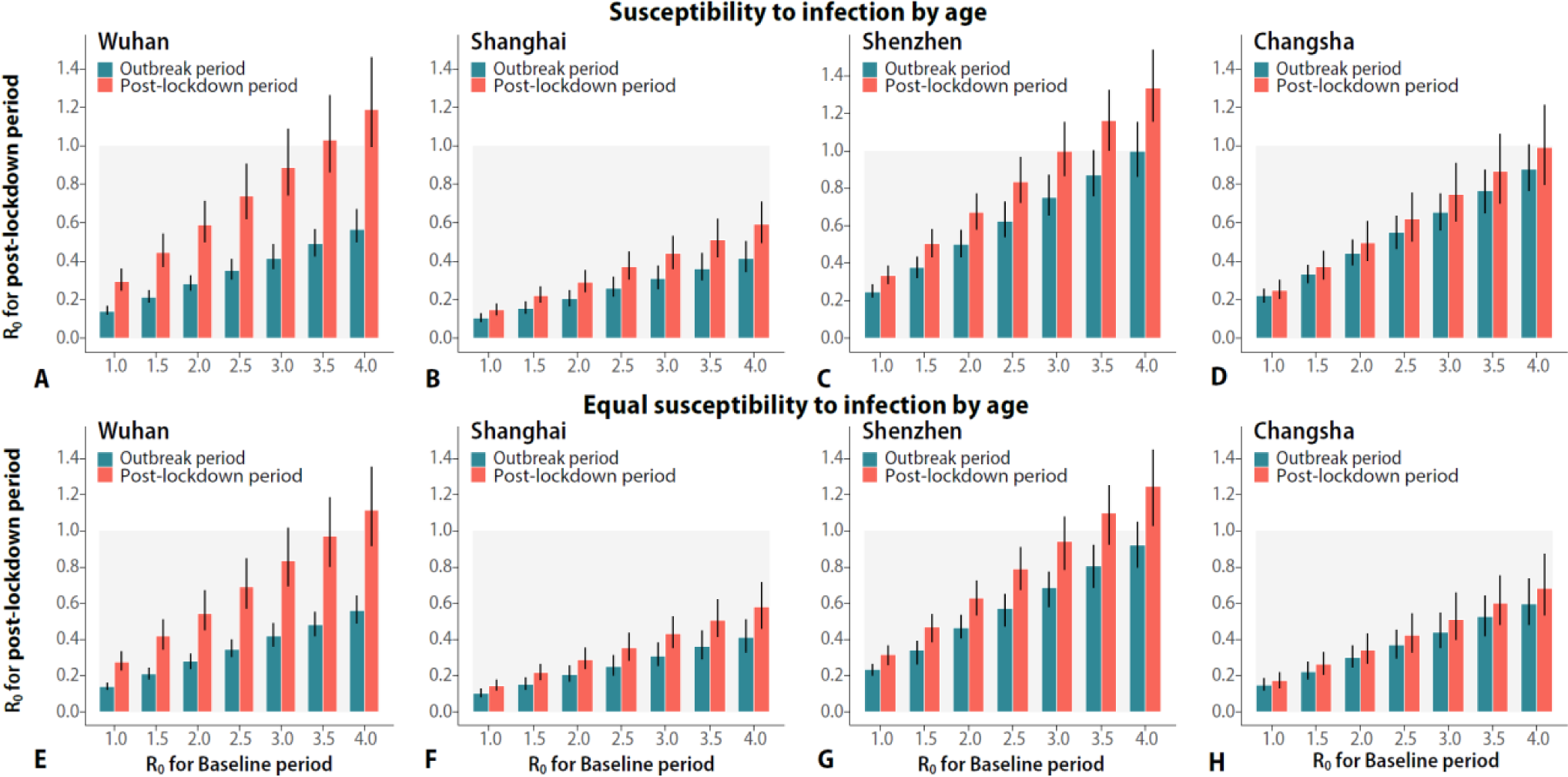
Effect of relaxing interventions on epidemic spread (assuming equal infectiousness by age). (**A**) Estimated R_0_ during the outbreak and post-lockdown periods (mean and 95% CI) as a function of baseline R_0_ (i.e., that derived by using the contact matrix estimated from the baseline period). The distribution of the transmission rate is estimated through the next-generation matrix approach by using 100 bootstrapped contact matrices for the baseline period to obtain the desired R_0_ values. We then use the estimated distribution of the transmission rate and the bootstrapped outbreak and post-lockdown contact matrices to estimate R_0_ for the outbreak period and the post-lockdown period, respectively. The 95% CIs account for uncertainty in the distribution of the transmission rate, mixing patterns, and susceptibility to infection by age. (**B-D**) Same as (A), but for Shanghai, Shenzhen, and Changsha. The figure includes both the scenario accounting for susceptibility to infection by age and the scenario where we assume that all individuals are equally susceptible to infection. (A-D) refer to the first scenario, and (**E-H**) refer to the second scenario.

When we considered contacts in the post-lockdown period, while keeping the same baseline disease transmissibility as in the pre-intervention period at an average value of R_0_=2.5, the increase in the number of contacts was not high enough to bring R_0_ above 1 in any location. The reproductive number exceeded the epidemic threshold in Wuhan (Fig. 3A) and Shenzhen (Fig. 3C) for baseline R_0_ above 3.0, and in Changsha (Fig. 3D) for baseline R_0_ above 3.5. The reproductive number in Shanghai did not reach the threshold even for baseline R_0_=4 (Fig. 3B). These findings are robust to assumptions about age differences in susceptibility to infection in Wuhan, Shanghai, and Shenzhen (Fig. 3E-3G). Considering age differences in susceptibility to infection has a marked effect in predicted dynamics in Changsha, where transmission can be interrupted for baseline R_0_ at least up to four (Fig. 3H). We performed sensitivity analyses regarding age differences in infectiousness; an average individual aged 0-14 years was assumed to be twice less infectious than one aged above 14 years old. The results are consistent with those reported here (Supplementary Material, Sec. 6.1). We also performed sensitivity analyses regarding possible compliance biases of self-reported contacts during the post-lockdown period. In this case, by assuming baseline R_0_= 2.5 and by making the extreme assumption that only half of the actual non-household contacts were reported by study participants, the reproduction number for post-lockdown period would still be below the threshold or slightly above it (Supplementary Material, Sec. 6.2).

### Impact of a possible return to pre-pandemic mixing patterns in schools, workplaces, and the community

We performed scenario analyses for Shanghai where we assume that contacts at school, in the workplace, and the community return to the levels reported during the pre-pandemic period. In this analysis, we set R_0_=2.5 to be in line with R_0_ measured in Wuhan before the start of the interventions (*16*) and age-specific susceptibility to infection as estimated in our previous study (*9*).

By assuming no contacts at school, we estimate that, in the absence of contact tracing and other control measures, 55% of workplace and 65% of community mixing could be resumed while keeping R_0_ below the epidemic threshold. In particular, a reduction of 80% of effective contacts in the community (corresponding to 1.2 contacts per day), would allow the work contacts to be resumed at 50% (Fig. 4A). Opening of senior high schools, even when considering as little as 10% of workplace and 10% of community contacts, could increase R_0_ above 1 (Fig. 4B). Broader school opening has higher effect on R_0_ (Fig.4C).

**Fig. 4.**
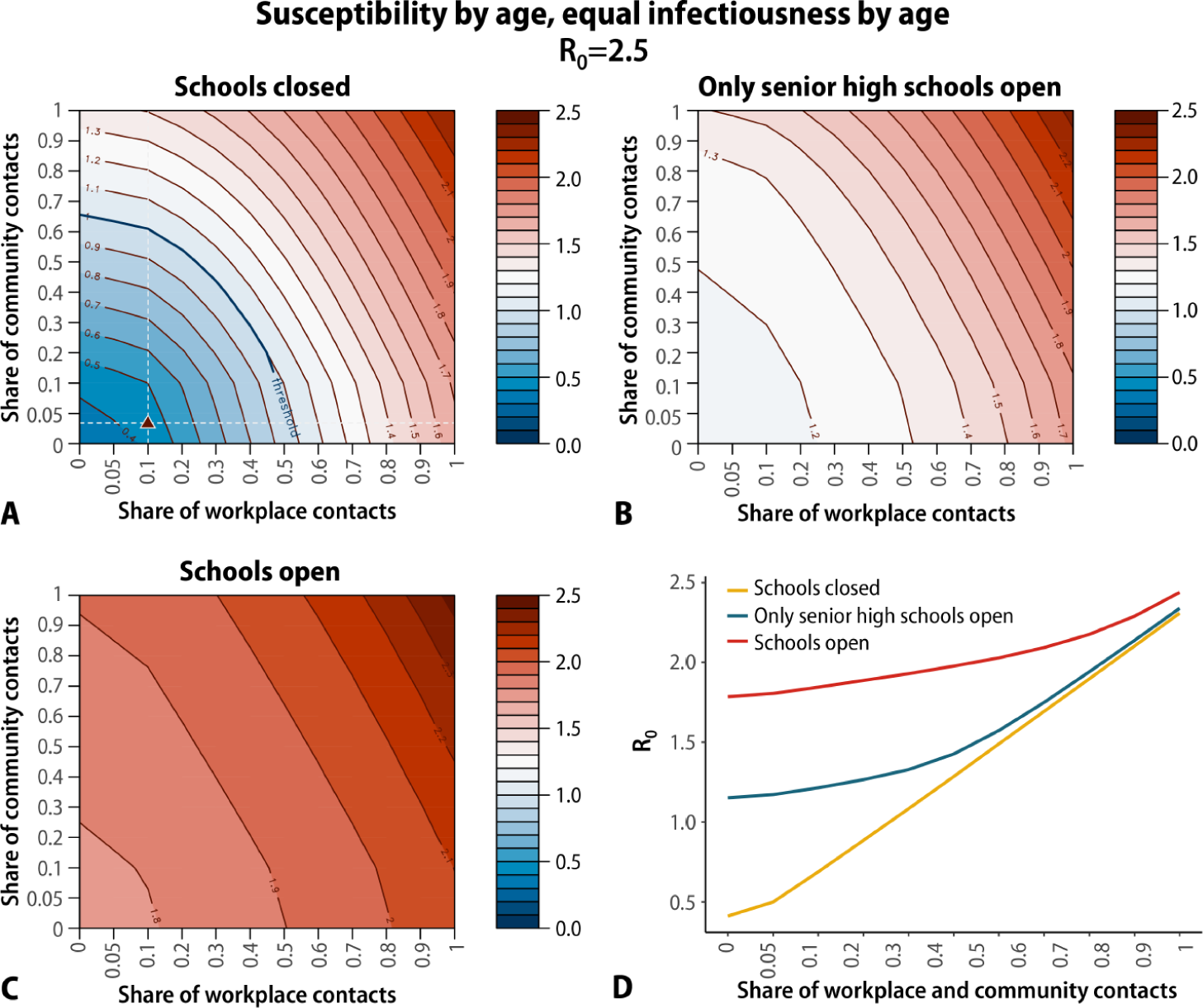
Estimated R_0_ for Shanghai under different assumptions on the number of contacts in workplaces, community, and schools. **A** Heatmap of the estimated mean value of R_0_ for a different combination of number of contacts in the workplaces and community under the assumption that all schools are closed. Share equal to 1 corresponds to the pre-pandemic contact pattern. The triangle corresponds to the measured share of contacts in the workplace and the community during the post-lockdown period. Baseline period R_0_ was set to 2.5, we considered age-specific susceptibility to infection as in (*9*), and no differences in infectiousness by age. R_0_ values are estimated through the next generation matrix approach. **B** Same as A, but assuming that contacts in senior high schools are as in the pre-pandemic period. **C** Same as A, but assuming that contacts in all schools (corresponding to the entire student population except for college students) are as in the pre-pandemic period. **D** Estimated mean value of R_0_ for different shares of workplace and community contacts under different levels of school closure.

It is important to highlight that the effect of school closure depends non-linearly on the share of other activities being resumed (Fig. 4D). In particular, if we compare R_0_ in an extreme scenario when nearly all workplace and community activities are halted (i.e., only 5% workplace and community contacts included, which is about the same level observed during the lockdown period – about 4%), we can see that school closure corresponds to a decrease of R_0_ from 1.78 to 0.41. On the contrary, in a scenario where workplace and community contacts return to their pre-pandemic level (i.e., 100%), school closure corresponds to a decrease of R_0_ of about 0.13. In other words, social contacts in schools only truly matter if other social mixing opportunities are hindered. In such context of greatly reduced contact patterns at work and in the community, school closure becomes determinant in controlling disease spread. When contacts in other social settings are moderate or back to normal, then school closure is less relevant as the infection can spread through other chains of transmission. This effect is non-linear and is captured by the change of the eigenvalues of the next generation matrix.

When we consider relaxing interventions simultaneously (e.g., school reopening, workplace resumption, resuming community activities), returning up to 34% of the pre-pandemic level of non-household contacts (corresponding to a mean of 7.8 contacts per day) can lead R_0_ above the threshold in Shanghai, with slightly lower proportions (26% to 30%, corresponding to 3.4 to 5.5 contacts per day) in Shenzhen, Changsha, and Wuhan (Supplementary Material, Sec. 9).

## Discussion

We have provided measures of the changes in mixing patterns linked to gradual relaxation of interventions in the Chinese cities of Wuhan, Shanghai, Shenzhen, and Changsha and their impact on the spread of SARS-CoV-2. We find that although the number of contacts has increased in the post-lockdown period in China, social behavior has only partially and slowly changed and mixing patterns have not returned to normal, including among adults. We estimate that the increase in mixing patterns one month after the Wuhan lockdown and after work resumed in the other three cities was not enough to sustain local transmission. In fact, the mixing patterns measured in this study were far from the pre-pandemic ones. We also estimate that the importance of school closure/reopening on SARS-CoV-2 control increases in a non-linear fashion as contacts in the other settings decrease. In particular, our findings support that closing schools was essential to keep the epidemic under control during the lockdown.

Several studies have shown a very marked decrease in the number of contacts between the pre-pandemic and lockdown periods(*9, 20-24*), in agreement with what we found here, which parallels a decrease in mobility patterns (*9, 25*). However, our study shows that the post-lockdown increase in mobility patterns is not a good proxy of the daily number of in contacts in the four study locations (Supplementary Material, Sec. 4). Indeed, the intra-city mobility in the analyzed cities has also been rebounding to pre-pandemic levels, while contacts in the same period are still very close to the ones estimated during the lockdown period. Several studies for different countries have shown a rebound in mobility after the lockdown (*26-29*). However, the much slower increase in the number of daily contacts that we estimated here could be location-specific. A similar pattern has been found in the US where mobility measured by mobile phone data has rebounded to pre-pandemic levels, but the number of contacts quantified by co-proximity is still at 50% or less of pre-pandemic levels (*30*). Whether these results can be generalized to other countries remain unclear.

To estimate how changes in contact patterns affect SARS-CoV-2 spread, we performed a model-based analysis. In particular, we estimated that the level of contacts recorded during the post-lockdown period in Spring 2020 are sufficient to keep the reproduction number below the epidemic threshold. On the contrary, in a purely theoretical scenario where all schools were open but parents could agree to stay at home, social distancing among adults would not be sufficient to prevent a COVID-19 outbreak. This highlights the need of considering additional case- and population-based interventions such as those in place in China since the end of the lockdown (e.g., 14-day quarantine in designated places for medical observation and testing for imported cases, use of face masks). However, it is important to stress that the population may spontaneously adjust its behavior, including avoidance of gatherings and adherence to hygiene measures, to limit their risk of SARS-CoV-2 infection while the pandemic is still spreading worldwide. These behavioral adaptations may partially explain why the number of contacts measured in this study in the post-lockdown period is still remarkably lower than before the pandemic.

This study is prone to the limitations pertaining to social contact surveys. The definition of contact relevant for SARS-CoV-2 transmission is still unclear. We used the classic definition of having a conversation or direct physical contact (*9*), which does not include contacts with surfaces (fomites transmission) that may have been possibly contaminated by infectious individuals. However, contacts with surfaces are hard to quantify, irrespective on the adopted methodology, and their contribution on SARS-CoV-2 transmission has yet to be clarified. The contact survey presented in this study is based on self-reported contacts. It can thus be affected by various biases, including recall bias and self-reporting bias. In particular, reported contacts for the baseline period in Wuhan, Shenzhen, and Changsha may be prone to recall bias as contacts were recorded retrospectively. This may explain why we observed a larger number of contacts during that period in Shanghai with respect to the other three locations; as well as less marked typical features of contact matrices by age (such as the presence of three main diagonals representing contacts among household members, a bottom-left corner showing contacts among students, and a central area showing contacts between workers) in this three locations with respect to Shanghai. Nonetheless, it is important to stress that the mean number of daily contacts estimated for the four locations during the pre-pandemic period are comparable to those obtained in other countries(*10, 31, 32*), ranging from 4.5 contacts per day in Japan(*32*) to 27 contacts per day in UK(*33*). Moreover, the age-stratified contact matrices presented in those studies share the same features as those presented here. Another possible bias is that survey participants may have felt pressure to minimize reported contacts that occurred during the post-lockdown period, given that social distancing policy has not been totally relaxed, even if the anonymity and confidentiality of the survey were emphasized. However, results are robust to inflating reported contacts outside of the home severalfold, suggesting that possible compliance and social acceptability biases linked to the post-lockdown period do not affect our main findings (Supplementary Material, Sec. 6.2).

The performed modeling analysis contains several approximations. The model does not consider explicitly symptomatic and asymptomatic individuals and possible differences in their infectiousness. The model assumes a homogeneous network of contacts (i.e., it does not account for the typical clustering of human populations) and thus the obtained results can be considered as upper bounds of SARS-CoV-2 transmissibility. The adopted transmission scheme is simple and does not explicitly account different levels of the severity pyramid, such as hospitalizations or deaths. Nonetheless, incidence and disease burden estimates are beyond the aim of the paper as the model was designed to provide a general picture of the effect of age-mixing patterns on the SARS-CoV-2 transmission rather than projections of the impact on the healthcare system. Finally, we would like to stress that here we do not necessarily endorse relaxing or reinstating social distancing policies in the context of COVID-19, but rather merely describe their impact on SARS-CoV-2 transmission based on the data collected in the four Chinese cities analyzed. Whether these findings can be generalized to other locations warrants further research. Future studies will be needed to assess the situation in China as of July 2020; in fact, over the period from May 15 to July 31, 2020, five outbreaks of local transmission have been reported for a total of 986 locally transmitted cases in mainland China.

Our study provides evidence that relaxing interventions in Wuhan, Shanghai, Shenzhen, and Changsha, and the resulting changes in human behavior, have slowly raised the number of daily contacts among adults. Moving forward, in order to prevent the resurgence of the epidemic, it will be particularly important to strengthen sanitization activities (e.g., washing hands, disinfection) and proactive social distancing measures (e.g., increased distance between individuals while in contact or use of a face mask) in the post-lockdown phase, along with large-scale testing and contact tracing(*34-37*). This is particularly important if schools are reopened(*38*). Researches should continue to focus on refining age-specific estimates of susceptibility to infection, infectiousness, and risk of severe disease, in conjunction with changes in mixing patterns, as these are instrumental to evaluate the impact of control strategies on SARS-CoV-2 transmission.

## Materials and Methods

### Design of the contact survey

To estimate changes in age-mixing patterns associated with relaxation of interventions against SARS-CoV-2, we conducted contact surveys in four cities. The design of the survey was similar to our previous work(*9-11*). Participants in Shenzhen and Changsha were asked to complete a questionnaire describing their contact behavior(*9, 12*) on three different days: i) a regular weekday between December 24, 2019 and December 30, 2019, before the COVID-19 outbreak was officially declared (baseline period); ii) a day between February 3 and February 9, 2020 when the COVID-19 epidemic peaked across China and stringent interventions were in place (outbreak period); and iii) the day before the telephone interview took place, corresponding to a period when interventions were being relaxed across China (post-lockdown period). A contact was defined as either a two-way conversation involving three or more words in the physical presence of another person, or a direct physical contact (e.g., a handshake). Participants in Wuhan and Shanghai were asked to complete the same questionnaire used for Shenzhen and Changsha, but to report only the contacts during the post-lockdown period. In fact, for the baseline and outbreak period in Wuhan and Shanghai, we relied on a survey conducted in 2017-2018 and another survey conducted in 2020 during the pandemic that followed the same design(*9, 10*). A more detailed description of the methodology is given in the Supplementary Material (Sec. 1-2).

### Estimation of contact patterns

The number of daily contacts (including physical and conversational contacts) was compared between the post-lockdown period and the outbreak period for each location using a weighted negative binomial regression, where the weight refers to the day type of contact diary (i.e., weekdays or weekends)(*9*). The same comparison was performed among four cities for the outbreak period and the post-lockdown period, respectively.

We defined 16 age groups (0-4 y, 5-9 y, 10-14 y, 15-19 y, 20-24 y, 25-29 y, 30-34 y, 35-39 y, 40-44 y, 45-49 y, 50-54 y, 55-59 y, 60-64 y, 65-69 y, 70-74 y, and 75 y and over) to build age-specific contact matrices. Contact matrices representing mixing patterns during a baseline pre-pandemic day (referred to as “baseline period contact matrix”), during the COVID-19 outbreak (referred to as “outbreak contact matrix”), and during the period of relaxation of interventions (referred to as “post-lockdown contact matrix”) were estimated for the four cities. Due to the sample size, for baseline and outbreak contact matrices in Wuhan, we used 14 age groups (last age group being 65+ years). To account for the uncertainty of the contact matrices and sample representativeness of contact matrix, we performed bootstrap sampling with replacement of survey participants weighted by the age distribution of the actual population in each location. For the post-lockdown period, we adjusted for weekdays and weekends (weight of 2/7 for diaries referring to Saturdays/Sundays, weight of 5/7 for diaries referring to Monday to Friday) (*9*).

### SARS-CoV-2 natural history and transmission

To explore how our empirical data can inform how to relax COVID-19 control strategies, we performed a modeling exercise. A key parameter regulating the dynamics of an epidemic is the basic reproduction number (R_0_), which corresponds to the average number of secondary cases generated by an index case in a fully susceptible population. We estimated the impact of relaxing interventions on R_0_, relying on the empirically estimated mixing patterns. We also considered age-specific estimates of susceptibility to infection presented in(*9*) and no differential infectiousness by age(*39*). We also consider scenarios where all age groups are equally susceptible by age and where children are twice less infectious than adults (Supplementary Material, Sec. 6.1). The mean time interval between two consecutive generations of cases was set to be 5.1 days, aligning with the mean of the serial interval reported by Zhang et al(*40*). Finally, we used the next generation matrix approach to quantify estimate R_0_(*41*) (Supplementary Material, Sec. 5).

### Simulation of hypothetical scenarios with alternative mixing patterns

We project how increasing school, workplace, and community contacts can affect COVID-19 spread in Shanghai. We considered several different scenarios, based on the baseline contact matrix and gradual implementation of interventions by social setting (i.e., household, school, workplace, and community). A similar analysis was performed for Shenzhen and Changsha (Supplementary Material, Sec. 8). No simulations were performed for Wuhan, as for this location we did not collected data by social setting for the baseline period(*9*). We finally used the same next generation matrix approach to project what degree of return to pre-pandemic contact could lead R_0_ above the epidemic threshold if the contacts outside the household are resumed simultaneously (Supplementary Material, Sec. 9).

## Supplementary Materials

Materials and Methods

Figures S1 – S34

Tables S1-S30

References (42-53)

## Data Availability

All data and code is available in the main text or the supplementary materials or it will be made available on GitHub and Zenodo upon acceptance.

